# Feasibility of collecting and processing of COVID-19 convalescent plasma for treatment of COVID-19 in Uganda

**DOI:** 10.1101/2020.10.29.20222067

**Authors:** Winters Muttamba, John Lusiba, Loryndah Olive Namakula, Pauline Byakika-Kibwika, Francis Ssali, Henry Ddungu, Levicatus Mugenyi, Noah Kiwanuka, Rogers Sekibira, Cissy Kityo, Dorothy Keyune, Susan Acana, Ambrose Musinguzi, Ayub Masasi, Joseph Byamugisha, David Mpanju, Walter Jack Musoki, Hellen Aanyu Tukamuhebwa, Fred Nakwagala, Bernard Sentalo Bagaya, Alex Kayongo, Ivan Kimuli, Rebecca Nantanda, Winceslaus Katagira, Esther Buregyeya, Rosemary Byanyima, Baterana Byarugaba, Trishul Siddharthan, Henry Mwebesa, Olaro Charles, Moses Lutaakome Joloba, William Bazeyo, Bruce Kirenga

**Affiliations:** Makerere University Lung Institute, Kampala, Uganda; Uganda Peoples Defense Forces Medical Services; Department of Medicine, Makerere University College of Health Sciences, Kampala Uganda; Joint Clinical Research Centre; Uganda Cancer Institute; School of Public Health, Makerere University College of Health Sciences, Kampala Uganda; Uganda Blood Transfusion Services; Mulago National Referral Hospital, Kampala, Uganda; Immunology and Molecular Biology, Makerere University College of Health Sciences, Kampala Uganda; Pulmonary and Critical Care, Johns Hopkins University, Baltimore, Maryland, USA; Ministry of Health, Republic of Uganda, Kampala, Uganda; Makerere University, Kampala, Uganda; Uganda Heart Institute, Kampala, Uganda

## Abstract

**Introduction:** Evidence that supports the use of COVID-19 convalescent plasma (CCP) for treatment of COVID-19 is increasingly emerging. However, very few African countries have undertaken the collection and processing of CCP. The aim of this study was to assess the feasibility of collecting and processing of CCP, in preparation for a randomized clinical trial of CCP for treatment of COVID-19 in Uganda.

**Methods:** In a cross-sectional study, persons with documented evidence of recovery from COVID-19 in Uganda were contacted and screened for blood donation via telephone calls. Those found eligible were asked to come to the blood donation centre for further screening and consent. Whole blood collection was undertaken from which plasma was processed. Plasma was tested for transfusion transmissible infections (TTIs) and anti-SARS CoV-2 antibody titers. SARS-CoV-2 testing was also done on nasopharyngeal swabs from the donors.

**Results:** 192 participants were contacted of whom 179 (93.2%) were eligible to donate. Of the 179 eligible, 23 (12.8%) were not willing to donate and reasons given included: having no time 7(30.4%), fear of being retained at the COVID-19 treatment center 10 (43.5%), fear of stigma in the community 1 (4.3%), phobia for donating blood 1 (4.3%), religious issues 1 (4.4%), lack of interest 2 (8.7%) and transport challenges 1 (4.3%). The median age was 30 years and females accounted for 3.7% of the donors. A total of 30 (18.5%) donors tested positive for different TTIs. Antibody titer testing demonstrated titers of more than 1:320 for all the 72 samples tested. Age greater than 46 years and female gender were associated with higher titers though not statistically significant.

**Conclusion:** CCP collection and processing is possible in Uganda. However, concerns about stigma and lack of time, interest or transport need to be addressed in order to maximize donations.

## Introduction

There have been over 41 million cases of COVID-19 reported, and more than one million deaths recorded (1). In Uganda, the first confirmed case of COVID-19 was reported on 21^st^ March 2020 (2). Up till mid-May, the local COVID-19 epidemic spread at a slow pace and mainly comprised of imported cases, majority of whom were asymptomatic. There were limited foci of transmission with no evidence of community transmission (2). From mid-May to early August, the local epidemic progressed to more foci and clusters of transmission. However, starting from mid-August, there has been a rapid rise in the number of cases detected daily and rapid progression to community transmission and increasing mortality. As of 5^th^ October 2020, up to 8,808 cases have been recorded with 81 deaths reported (3).

Control of COVID-19 in Uganda has been mainly through the non-pharmacologic measures adopted from the recommendations of the World Health Organization (WHO). These measures include use of face masks, social distancing and hand washing or sanitization using alcohol containing sanitizers (4). With no vaccines for COVID-19 available, several repurposed and new drugs have been reported in compassionate use and small trials with mixed benefit (5).

Evidence is emerging to support the use of COVID-19 convalescent plasma (CCP) for treatment of COVID-19 especially among patients with severe and critical forms of disease (6–10). Administration of CCP has been found to be safe and associated with clinical, radiological and laboratory improvements as well as reduction in mortality (6,11–14). However, some studies found no benefit of CCP with regard to reducing mortality and or length of hospital stay, improving the day 15 disease free severity or shortening the time to clinical improvement(15,16). One study by Li et al was terminated early due to inability to reach the targeted sample size. Given the mixed and inconsistent nature of findings of CCP use, there is need for more rigorous studies to assess the efficacy of CCP in treatment of COVID 19.

To-date, there are few African countries that have undertaken the collection and processing of CCP (17). According to the international society of blood transfusion (ISBT) document library, only South Africa is conducting a CCP trial while Ghana has posted guidelines for collecting, processing, storage and distribution of CCP (18). The slow embracement of CCP is probably due to pre-COVID-19 pandemic challenges to blood transfusion services such as reliance on whole blood transfusions, widespread unavailability of blood component production technology, erratic power supply, inadequate storage capacity, transportation challenges, clinicians’ inexperience in the appropriate use of blood components, and limited financial resources (17).

With more than 4700 individuals recovering from COVID in Uganda (3), the increasing spread of the virus in the community and occurrence of severe and critical cases, we undertook a study to assess the feasibility of collecting, processing and storing CCP, in preparation for a randomized clinical trial of CCP for treatment of COVID-19 in Uganda.

## Methods

### Design and study site

We undertook a cross sectional study of individuals that had been diagnosed with COVID-19 and treated in Uganda, and had evidence of recovery (defined as two negative PCR tests performed at least 24 hours apart). The work was undertaken at Mulago National Specialized Hospital, where donor assessment and blood collection were performed. Mulago National Specialized hospital, located in the capital Kampala is the biggest referral hospital in Uganda and offers super specialized services. Whole blood processing to obtain plasma was conducted at the Uganda Blood Transfusion Service (UBTS) blood bank situated 2.5 kms away from the donation centre. UBTS is mandated to provide blood services in the country through donation, processing, storage and distribution of processed blood to all health facilities in the country. As an extra layer of safety for the donated plasma, the plasma was tested for presence of SARS-CoV-2 virus using the RT PCR assay at the Department of Immunology and Molecular Biology, Makerere University located within the site of the donation. Additional samples were stored in a biobank at the same laboratory.

### Participants

We targeted all individuals with documented recovery from COVID-19 infection who had been managed and discharged at a designated COVID-19 treatment hospital in Uganda. To be eligible, the donor had to provide written informed consent, have documented evidence of SAR-CoV-2 infection by reverse transcriptase polymerase chain reaction (RT-PCR) test and recovery defined as two negative RT-PCR tests performed at least 24 hours apart, be at least 18 years old and meet all criteria for blood donation as set by Uganda Blood Transfusion Services (UBTS). The UBTS criteria include: age between 17-60 years, weight ≥50Kgs, pulse rate of 60-100 beats/minute, temperature of 37±0.4°C, hemoglobin level of 12.5-16g/dl for females and 13.5-17g/dl for males, and last blood donation not less than 3 months and 4 months for males and females respectively. We excluded females with previous history of blood transfusion and/or pregnancy as well as participants who had documented evidence of a HIV positive status.

### Study Procedures

#### Donor screening and donation

Administrative clearance was obtained from the Ministry of Health to access a list of recovered individuals from the COVID-19 treatment hospitals in Uganda. Initial screening for blood donation eligibility was performed by telephone. Screening covered questions on recovery from COVID, age, weight, pregnancy history for females, days since diagnosis, comorbid history and whether the individual would be willing to donate. Eligible and willing donors were invited to the donation centre to complete further assessment for eligibility using the study eligibility criteria and the Uganda blood transfusion screening criteria for community donations. Weight was measured using a bathroom scale (SECA) and blood hemoglobin measured using a hemochromax machine. Up to 450mls of whole blood was collected in a quadruple bag using an automated bio mixer and transported to the blood bank in a cool box at 2-10°C within 8 hrs of collection. Donors were requested to return to the donation centre for their blood grouping results. At this visit, those that had TTIs were counselled and linked to care.

#### Plasma separation and storage

The LUXOmatic V2 blood separation device(19) was used for automatic extraction of the convalescent plasma. The separator uses a colormatic sensor system for best differentiation between red cells, platelets and plasma. Blood component separation of 2 blood bags could be done simultaneously with high quality of plasma component due to the sensitive sensor system. Up to 200mls of CCP was obtained from each 450mls unit of whole blood, labeled and immediately placed on flat in the freezer at –18°C, in a designated CCP shelf for 30 minutes, then stored in a deep freezer at –80°C. All processed CCP units were clearly labelled with an Investigation New product (IND) label. Three 2mls aliquots of plasma were obtained in cryo-vials and stored to permit retrospective determination of the characteristics of an effective product and future investigations.

#### Blood grouping and pretesting

The Immucor blood grouping Galileo system was used to perform ABO and Rh blood grouping while screening for transfusion transmissible infections (TTIs) was done using the Abbott Architect i2000 technology. Positive units for TTIs were repeated on Fortress ELISA as per the blood bank testing algorithm. We tested plasma for presence of SARS CoV-2 RNA using the ALTONA RealStar SARS CoV-2 RT-PCR kit following the manufacturer’s instructions (20).

#### Biological sample banking

We collected blood samples, processed them and bio banked them for future use. Samples bio banked included PBMCs, Serum and Plasma.

#### SAR-CoV2 Antibody titer testing

Antibody titers were determined using the Acro Biosystems Anti-SARS CoV-2 antibody IgG titer Serological ELISA Assay kit (Spike protein RBD)(21) following the manufacturer’s instructions. The resultant IgG titers (nanograms/milliliter) was converted to arbitrary units by dividing with the concentration (nanograms/milliliter) that corresponded with three times the average optical density (OD) of the negative control samples.

### Statistical analysis

Participants’ characteristics were described using proportions, means (standard deviation-SD), and medians (inter-quartile range-IQR) to describe the. Comparisons of characteristics were done using Chi-square or Fisher’s exact tests for categorical variables, and *t*-tests for continuous ones. All analyses were done using STATA® version 15.

### Ethical procedures

The study was conducted in accordance with the principles of the Declaration of Helsinki and in accordance with the principles of Good Clinical Practice (GCP) as laid down by the ICH topic E6 (Note for Guidance on GCP). Before initiating the study, ethical approval was obtained from the Mulago Hospital Research and Ethics Committee (MHREC 1872), as well as the Uganda National Council for Science and Technology (HS684ES). All participants gave informed written consent, and confidentiality of participants was through use of patient identification numbers instead of names on all study documents and samples. All patient information was kept secure and was only available to the study staff, regulators, and ethics committees.

## Results

### Donor screening

The outcome of donor screening is summarized in figure 1. Up to 192 participants were approached (8 females, 184 males) and of these, 179 (93.2%) were eligible to donate. The reasons for ineligibility for the 13 (6.8%) participants included: hypertension (23.1%), diabetes (15.4%), hypertension and diabetes (7.7%), HIV (7.7%), HIV and diabetes (7.7%), age below 18 years (7.7%), underweight (7.7%) and female with a history of pregnancy (23.1%). Of the 179 eligible to donate, 23 (12.8%) were not willing to donate. Reasons given for their unwillingness to donate included: having no time 7(30.4%), fear of being retained at the COVID-19 treatment center 10(43.5%), fear of stigma in the community 1(4.3%), phobia for donating blood 1(4.3%), religious issues 1(4.4%), lack of interest 2(8.7%) and transport challenges 1(4.3%). All eligible individuals were invited to the donation centre for further screening and donation, of whom 123 (78.8%) presented for additional screening. A further 63 recovered COVID-19 individuals were self-referred, increasing the total number of participants screened to 186. Of the 186 who came to the donation center, 24(12.9%) were ineligible at time of enrollment due to: high blood pressure (54.2%), HIV positive status (8.3%), HIV positive status and hypertension (4.2%), high temperature (4.2%), previous donation within one month (4.2%), underweight (8.3%), heart disease (4.2%), donation phobia (4.2%), on treatment for peptic ulcer disease (4.2%) and work pressures (4.2%).

**Figure 1:**
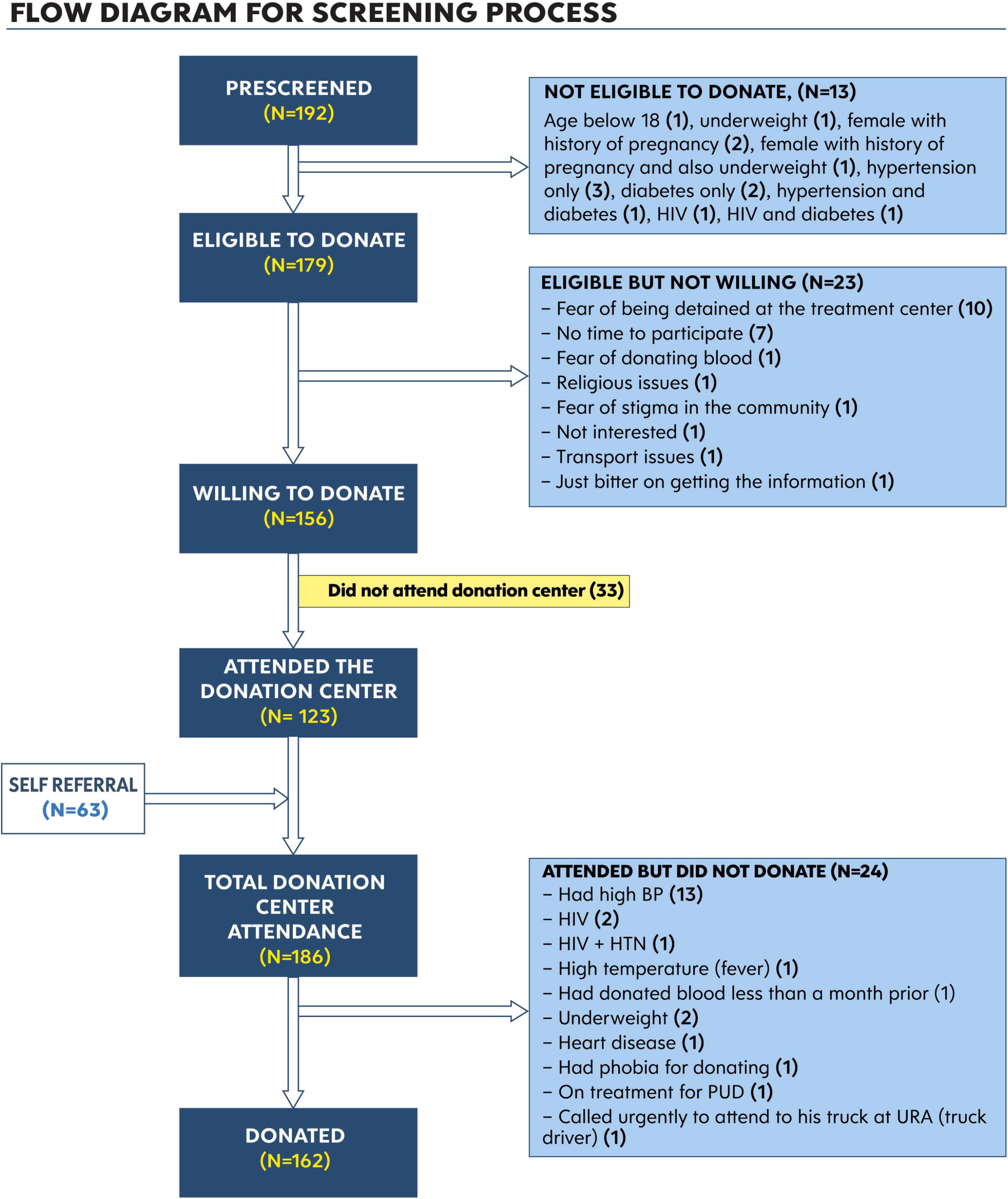
Flow diagram for screening process

### Social demographic characteristics of the donors

Table 1 below summarizes the social demographic characteristics of the donors. The median age was 30 years and females accounted for were 3.7% of the donors. Majority (64.2%) had attained secondary level of education. Over half (56.8%) had been previously managed at a regional referral hospital. The furthest distance travelled to the donation centre was 498km and average distance travelled was 161km.

**Table 1:**
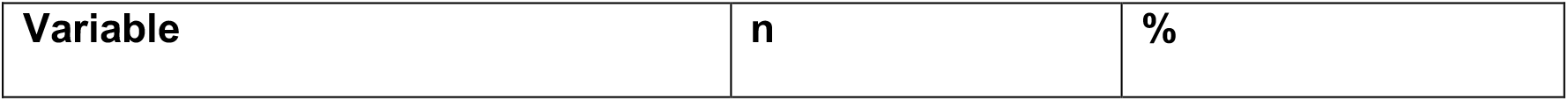
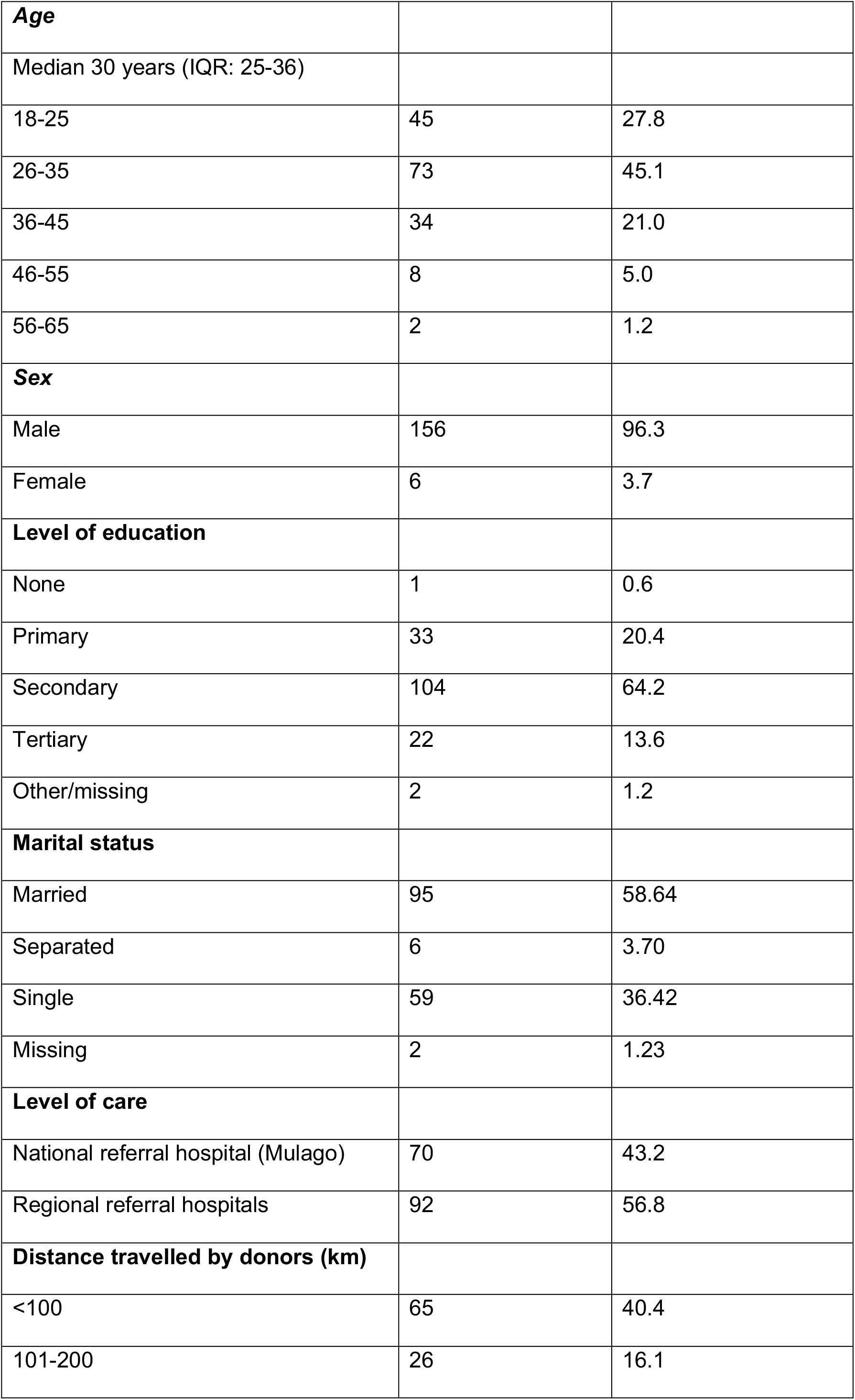
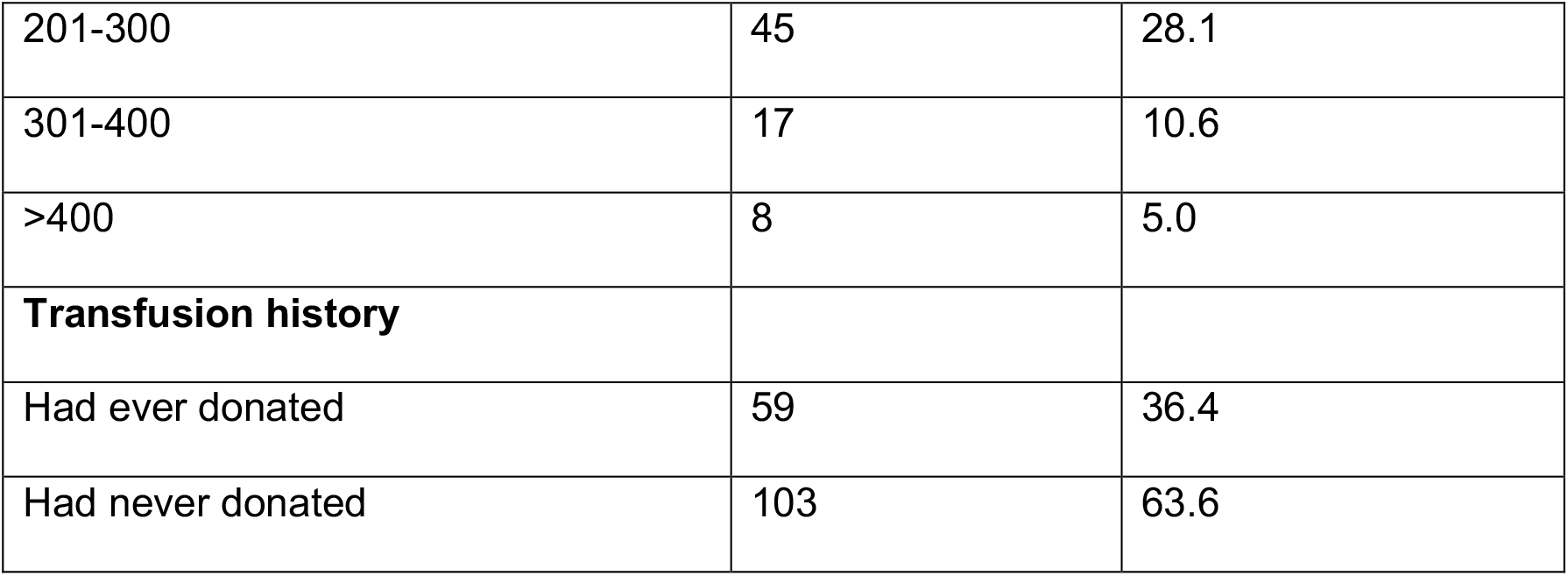
Characteristics of the donors (N=162)

| Variable | n | % |
| --- | --- | --- |
| **Age** |  |  |
| Median 30 years (IQR: 25-36) |  |  |
| 18-25 | 45 | 27.8 |
| 26-35 | 73 | 45.1 |
| 36-45 | 34 | 21.0 |
| 46-55 | 8 | 5.0 |
| 56-65 | 2 | 1.2 |
| **Sex** |  |  |
| Male | 156 | 96.3 |
| Female | 6 | 3.7 |
| **Level of education** |  |  |
| None | 1 | 0.6 |
| Primary | 33 | 20.4 |
| Secondary | 104 | 64.2 |
| Tertiary | 22 | 13.6 |
| Other/missing | 2 | 1.2 |
| **Marital status** |  |  |
| Married | 95 | 58.64 |
| Separated | 6 | 3.70 |
| Single | 59 | 36.42 |
| Missing | 2 | 1.23 |
| **Level of care** |  |  |
| National referral hospital (Mulago) | 70 | 43.2 |
| Regional referral hospitals | 92 | 56.8 |
| **Distance travelled by donors (km)** |  |  |
| <100 | 65 | 40.4 |
| 101-200 | 26 | 16.1 |
| 201-300 | 45 | 28.1 |
| 301-400 | 17 | 10.6 |
| >400 | 8 | 5.0 |
| **Transfusion history** |  |  |
| Had ever donated | 59 | 36.4 |
| Had never donated | 103 | 63.6 |

### Transfusion transmitted infections (TTI)

A total of 30(18.5%) donors tested positive for different TTIs. The prevalence of different TTIs were as follows: HIV (4, 2.5%), Hepatitis B (8, 5%), Hepatitis C (14, 8.8%) and Syphilis (4, 2.5%). From all the collected plasma, 127 samples were found viable for donation.

### SAR-CoV2 Antibody titers

All the CCP samples tested negative for SARS-Cov-2 by RT-PCR. Antibody titer testing demonstrated titers of more than 1:320 for all the 72 samples tested. Figure 2 demonstrates the antibody titers classified by age group, gender, HIV status and region of the donors while figure 3 demonstrates the antibody titers by number of days since first positive COVID result or days since admission. Age greater than 46 years and female gender were associated with higher titers though this was not statistically significant.

**Figure 2:**
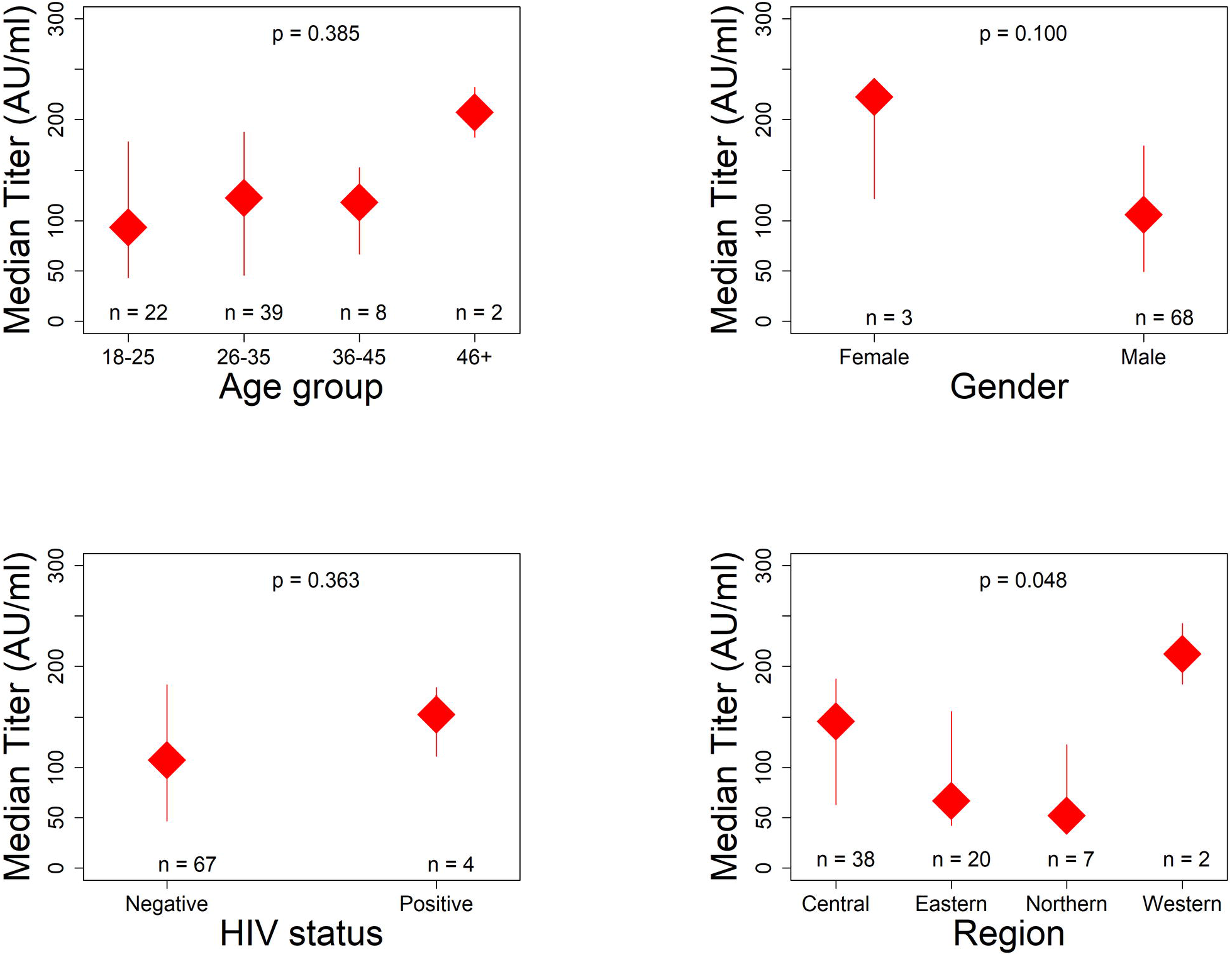
Antibody titers (AU/mL) classified by age group, gender and region of residence of donors

**Figure 3:**
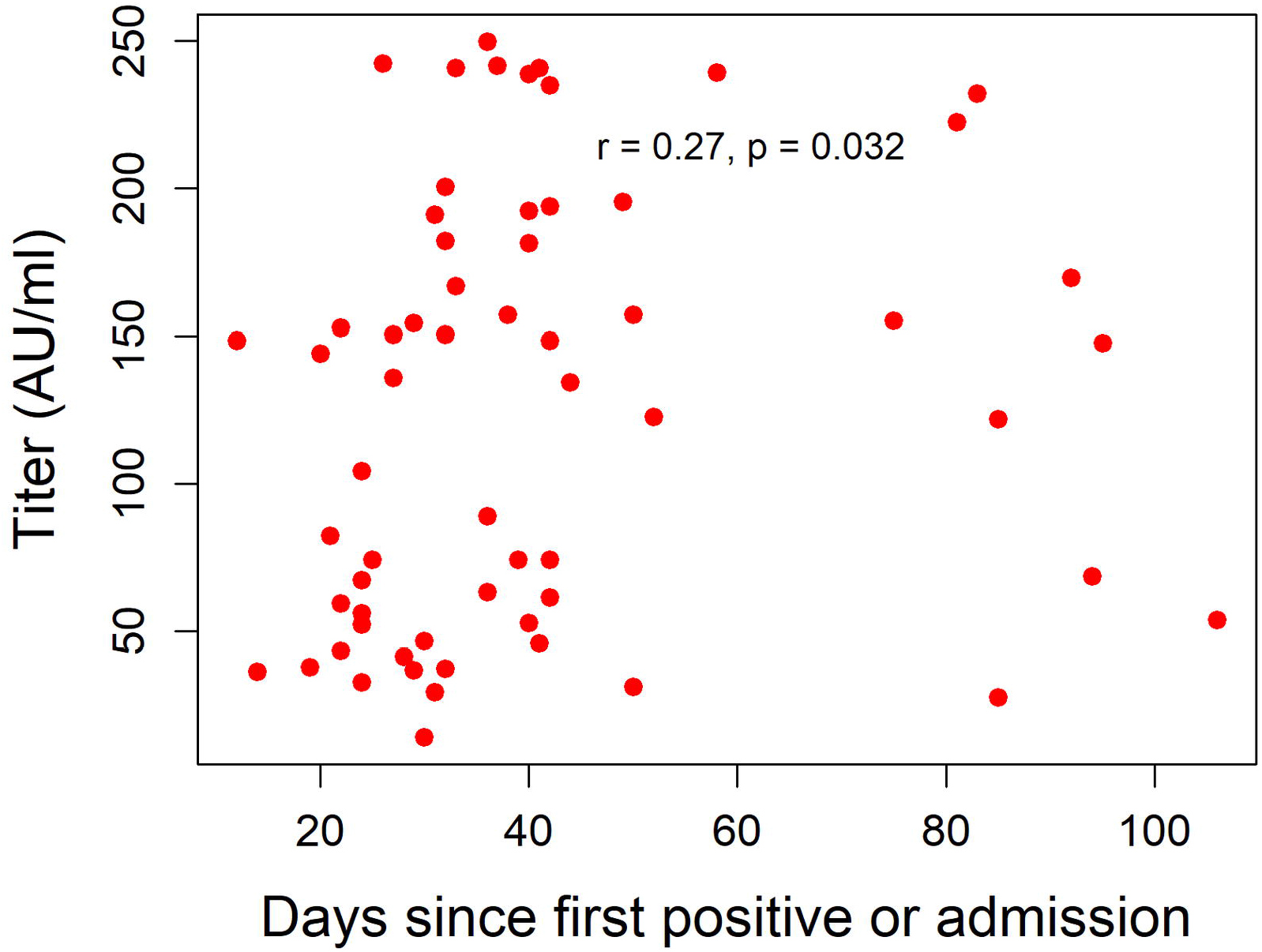
Antibody titers (AU/mL) by days since positive COVID test or admission

### Bio specimens

Table 2 below summarizes the type and quantities of the samples collected. Blood samples collected from the donors have been processed and bio banked.

**Table 2:** Summary of samples processed and bio banked

| Sample name | No. of participants | Vials per patient | Number of vials |
| --- | --- | --- | --- |
| Plasma | 162 | 4 | 545 |
| Serum | 52 | 2 | 104 |
| PBMC | 162 | 4 | 648 |

## Discussion

We aimed to assess the feasibility of collecting and processing CCP, in preparation for a randomized clinical trial of CCP for treatment of COVID-19 in Uganda. We have demonstrated that collection and processing of CCP is feasible and that all the donors have adequate antibody levels.

More than three quarters (93.2%) of the recovered individuals were found eligible to donate, with up to 87.2% willing to donate. The proportion eligible to donate in our study is higher than was found by Li et al in a pilot program (22). The difference could be attributed to their small sample size and level of screening where the 93.2% in our study was due to telephonic prescreening. The numbers would be expected to drop after following onsite/physical screening as individuals initially cleared as eligible may fail eligibility while at the donation site. It may additionally be due to younger demographics in the present study.

Donor eligibility and recruitment are an integral part of a successful CCP collection program. We instituted an eligibility criteria as has been recommended previously (23). The eligibility included evidence of a COVID 19 positive test and subsequent proof of recovery as evidenced by a discharge certificate at presentation to the donation centre. Donors for CCP need to satisfy other eligibility for routine donations (24), and thus to ensure deferrals to a minimum, a trained psychologist administered a pre-screening questionnaire by telephone, asking for obvious morbid conditions like HIV, Hepatitis, Hypertension, Diabetes, Syphilis among others.

In this study, 12.8% of the recovered individuals screened and found eligible to donate opted out for various reasons including stigma. Stigma has previously been identified to affect treatment seeking and also affect the recovered individuals (25–27).

Transfusion transmissible infections including COVID-19 have been highlighted as a potential challenge to CCP administration especially in low and medium income countries (18). There is no evidence for transmission of COVID-19 through blood donation (28); however, we ensured the donors were clinically and virally free of SARS-CoV-2. We did SARS-CoV-2 testing on the nasopharyngeal swabs collected from the donors and further did SARS-CoV-2 testing on the plasma and found no RNA in plasma or swabs of the donors. The UBTS did further screening for transfusion transmissible infections (HIV, Hepatitis C, Hepatitis B and Syphilis). It has been previously found out that the prevalence of TTIs tends to be lower among repeat donors as repeat donations select healthier individuals (24).

This study will inform an ongoing clinical trial assessing the safety and efficacy of CCP. To be able to execute this, antibody titer levels will be key. Analysis on 72 donors revealed they all had sufficient antibody titers in excess of 1:320, consistent with findings from a pilot program in Wuhan China where all the donors had sufficient antibody titers (22). The high levels of antibodies could explain why majority of the Ugandan patients had mild forms of disease and thus were able to mount a good humoral immune response. The high antibody titers could also be explained by the fact that most of the donors were included 28 days post diagnosis. The 28 day period of convalescence has been associated with a high antibody level (29).

Specimen biobanking is critical in pandemic response, and biospecimen issues have long been appreciated in emergency infectious disease control (30). In this feasibility study, we have been able to collect, process and store samples from recovered COVID 19 individuals for future research. We shall be able to link this to clinical data and answer any other emerging biomedical questions including monoclonal antibody manufacture.

## Limitations

Despite demonstrable feasibility of the collection and processing of CCP, the study had limitations including inability to do apheresis as had originally been planned. This would have allowed us to do more frequent donations from the same donors. We were unable to collect all the needed data from the participants who didn’t donate plasma. This would have given us an opportunity to compare the successful donors and the those that were screened out. We were also unable to do pathogen inactivation as is usually recommended for CCP.

## Conclusion

In our experience, CCP collection, processing and storage is possible in Uganda and should be possible in most low and middle income countries (LMICs) with functional national blood transfusion services. However, concerns about stigma and lack of time, interest or transport need to be addressed in order to maximize donations.

## Data Availability

Dataset analyzed during this study can be made available to all interested researchers upon reasonable request directed to the corresponding author

## Acknowledgements

We thank the Government of Uganda for funding the project through the Makerere University Research and Innovations Fund. We acknowledge Ministry of Health’s support and the support of all the implanting partners (Mulago national referral hospital, Uganda Peoples Defense Forces Medical Services, Joint Clinical Research Centre, Uganda Blood Transfusion Services). We thank the study participants and the health workers that collected the data as well as Ronald Kusolo and Priscilla Eroju who entered the data into the database.

